# Exact solution of infection dynamics with gamma distribution of generation intervals

**DOI:** 10.1101/2020.10.17.20214262

**Authors:** Alexei Vazquez

## Abstract

Infectious disease outbreaks are expected to grow exponentially in time but their initial dynamics is less known. Here I derive analytical expressions for the infectious disease dynamics with a gamma distribution of generation intervals. Excluding the exponential distribution, the outbreak grows as a power law at short times. At long times the dynamics is exponential with a growth rate determined by the basic reproductive number and the parameters of the generation interval distribution. These analytical expressions can be deployed to do better estimates of infectious disease parameters.

## I. INTRODUCTION

Infectious diseases can spread to a significant fraction of the population causing an epidemic. The chance for that to happen is determined by the infectious dynamics within each individual and the disease transmission between individuals. The within individual dynamics is reflected on the generation interval, the time interval between a primary case being infected to the disease transmission to a secondary case. The transmission between individuals is reflected in the reproductive number, the average number of secondary infectious caused by a primary case.

The networks underlying proximity and sexual transmission of infectious diseases have wide connectivity distributions and exhibit the small-world property [1, 2]. In these networks the basic reproductive number is proportional to the ratio between the second and first moments of the connectivity distribution [3–6], which can be very large. Less attention has been given to the shape of the generation interval distribution.

## II. SIR MODEL

Before entering the main calculations, let’s have a look at standard models of infectious disease dynamics. In the susceptible, infected and removed (SIR) model, susceptible individuals get infected at rate *β* and infected individuals get removed at a rate *γ*. The basic reproductive number, the average number of new infections generated by a single infected individual, is given by

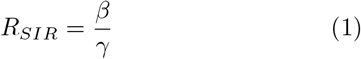

The dynamics of the infected individuals at the population level is given by

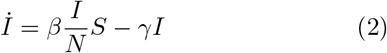

where *S* and *I* are the number of susceptible and infected individuals and *N* is the population size. For short times *S≈ N* and *I≪N*. In this limit we can integrate equation (7) obtaining

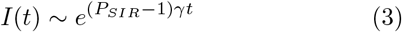

where

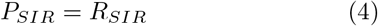

is the population reproductive number of the SIR model. According to the SIR model, the basic and population reproductive numbers are identical.

## III. SEIR MODEL

In the SEIR model susceptible individuals get infected at rate *β* without becoming infectious (exposed), exposed individuals become infectious at a rate *α* and infectious individuals get removed at a rate *γ*. The infected compartment is split into exposed, with number *E*, and infectious, with number *I*. Note that once an individual become infectious it behaves exactly as an infected individual in the SIR model. Therefore we obtain the same basic reproductive number

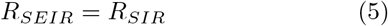

The dynamics of exposed/infectious individuals reads

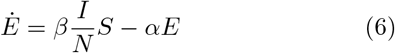

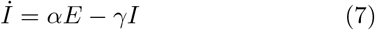

For short times *S ≈ N* and *I ≪N*. In this limit we can integrate equation (7) using an eigenvalue approach. Focusing on the largest eigenvalue, we obtain

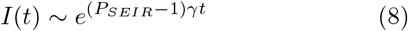

Where

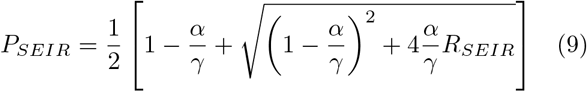

is the population reproductive number of the SEIR model. The introduction of the exposed compartment carries as a consequence a discrepancy between the local and population reproductive numbers, as previously noted [7].

## IV. BRANCHING PROCESS APPROACH

The SIR and SEIR models neglect key aspects of real populations. First, there is a variable contact rate across the population, which translates to a variable rate of disease transmission across infectious individuals. Second, while the SEIR model accounts for the incubation period of infectious diseases, it is still too restrictive. Using well stablished mathematics from the theory of age-dependent branching processes, I have previously calculated the expected number of infected individuals of infectious disease outbreaks on heterogeneous populations and any given time interval distribution [8, 9]. However, the applications were limited to an exponential distribution of generation intervals. Here I use this formalism to calculate the population reproductive number for a broader class of generation interval distributions.

The branching process approach maps contact processes mediating disease transmission into causal trees of disease transmission. The average reproductive number of patient zero, the expected number of secondary cases generated by patient zero, is given by

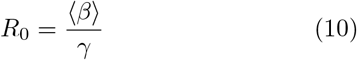

where ⟨*β*⟩ is the average infectious contact rate in the population and *γ* is the rate of recovery from the infection. For infected cases other than patient zero one needs to take into account that the disease spreading biases the disease transmission to individuals with a higher contact rate. The patient zero can be thought as an individual selected at random from the population. Any other infected individual will not be selected at random, but it will be found with a probability proportional to its contact rate: *β*/*N* ⟨*β*⟩, where *N* is the population size. Once infected, the individual found by contact will engage in new contacts at a rate *β*. Therefore, the average reproductive number of patients other than patient zero is

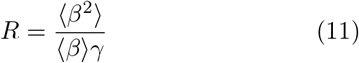

A similar distinction of reproductive numbers is made when considering spreading dynamics in static networks [3, 4]. In a static network the connectivity between agents is fixed, but agents have a variable number of neighbours, also known as degree and denoted by *k*. In that case the reproductive number of patient zero is proportional to the average degree ⟨*k*⟩. Furthermore, under the annealed or mean-field network approximation, the reproductive number of any agent other than patient zero is proportional to ⟨*k*^2^⟩ / ⟨*k*⟩ [3] or ⟨*k*(*k* − 1)⟩ / ⟨*k*⟩ [4], depending on the infectious model.

*R*_0_ gives the average number of infectious at the first generation, those generated by patient zero. *R*_0_*R* gives the average number of infections at the second generation and *R*_0_*R*^*d−*1^ gives the average number of infections at the *d* generation. The actual time when an infected case at generation *d* becomes infected equals the sum of *d* generation intervals and it has a probability density function *g*^*\* d*^(*t*), where the symbol *\** denotes convolution. Therefore, the average number of new infected individuals at time *t* is given by

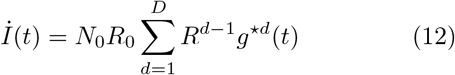

where *N*_0_ is the number of patients zero and *D* is the maximum generation, when the disease transmission ends. When *D* is small, due to lockdown for example, the spreading dynamics follows a polynomial or power-law growth [8, 10]. Here, I focus on the case *D → ∞*, or vmore precisely *t ≪D∫dtg*(*t*)*t*. In this limit

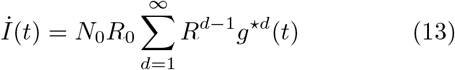

## V. GENERATION INTERVAL DISTRIBUTION

Now we will focus on the shape of the generation interval distribution. In the SIR model an infected individual is infectious right from the time it becomes infected until removed. Let *t*_*R*_ be a specific realization of the removal time. Since removal takes place at a rate *γ, t*_*R*_ has the exponential probability density function *γe*^*−γt*^. Assuming that each individual has a time-independent contact rate, the generation intervals will be uniformly distributed between 0 and the recovery time, resulting in the same exponential distribution of generation intervals

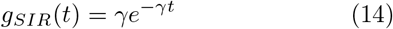

In the SEIR model the generation interval is decomposed into the sum *t*_*S*_ = *t*_*I*_ + *t*_*R*_, where *t*_*I*_ is the time interval from exposed to infectious and *t*_*R*_ is the removal time. Since the transition from exposed to infectious takes place at a constant rate *α*, then *t*_*I*_ has the exponential probability density function *αe*^*−αt*^. Therefore

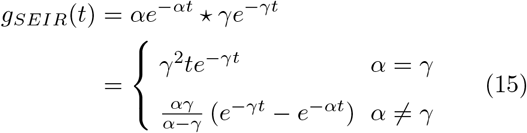

There are two key differences between the generation interval distributions for the SIR and SEIR model. First, the mode for *g*_*SIR*_(*t*) is zero and that for *g*_*SEIR*_(*t*) is non-zero. Second, around *t* = 0, *g*_*SIR*_(*t*) *∼ γ*, while *g*_*SEIR*_(*t*) *∼αγt*. Both differences are a consequence of introducing an incubation state between being infected and becoming infectious.

The case *α≠ γ* in (15) makes the calculation of convolutions difficult. A more suitable functional form is the gamma distribution

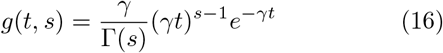

where *s ≥*1 is a shape parameter quantifying the convexity around *t* = 0 (Fig. 1). The case *g*(*t*, 1) corresponds with an exponential distribution, as in the SIR model (14). For *s* = 2 we obtain the case *α* = *γ* of the SEIR model (15). More generally, the values of *s* and *γ* could be derived from the fitting to empirical data. For example, for COVID-19, the inspection of inferred generation interval distributions suggest *γ >* 1 [11] and a fitting to a gamma distribution resulted in *γ* = 2.5 [12]. Besides covering many possible scenarios, the gamma distribution is amenable to convolution. Using the Laplace transform method one obtains

**FIG. 1.**
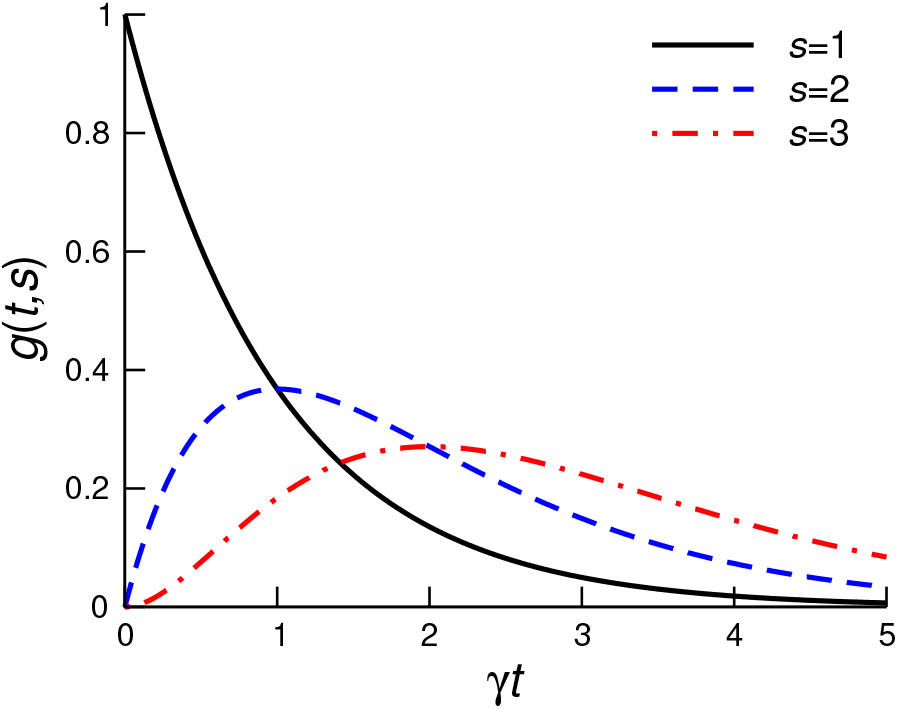
Gamma distribution with shape parameters *s*.

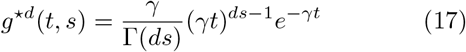

The convolution of a gamma distribution is itself a gamma distribution, with the exponent scaled by the order of the convolution (*d → ds*).

## IV. EXPECTED NUMBER OF INFECTIONS

Coming back to the outbreak dynamics, substituting (17) into (13) we obtain

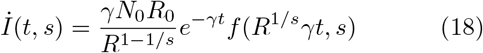

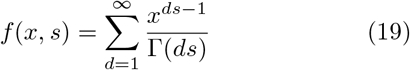

Equations (18)-(19) provide a series representation for the expected number of new infections. For short times, neglecting *d >* 1 terms we obtain (*R*^1*/s*^*γt ≪*1)

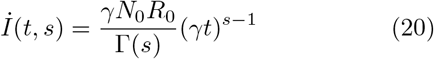

The case *s* = 1 is not representative. For *s* = 1 we have *İ*(*t, s*) *∼* 1, while *İ*(*t, s*) *∼ t*^*s−*1^ for *s >* 1. That is, except for the exponential distribution, the outbreak grows as a power law *t*^*s−*1^ for short times.

For *s* = 1 equation (19) is the series expansion of the exponential

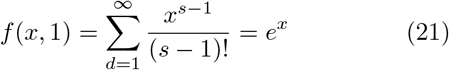

For *s* = 2 the series expansion of the hyperbolic sine

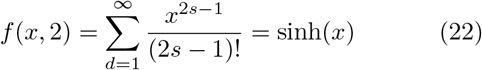

For *s* = 4 the series of sinh(*x*) minus sin(*x*)

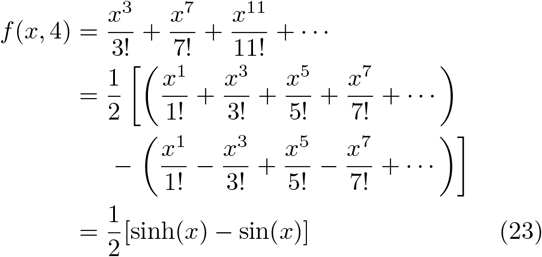

Using this analytical representations we uncover the full impact of the time generation distribution on the infection dynamics (Fig. 2). With increasing *s* there is an increase in the convexity of *f* (*x, s*) around *x* = 0 and the curves shift to the right. In contrast, the asymptotic behaviour for *x*≫ 1 seems to be the same. In the following I show that this is indeed the case.

**FIG. 2.**
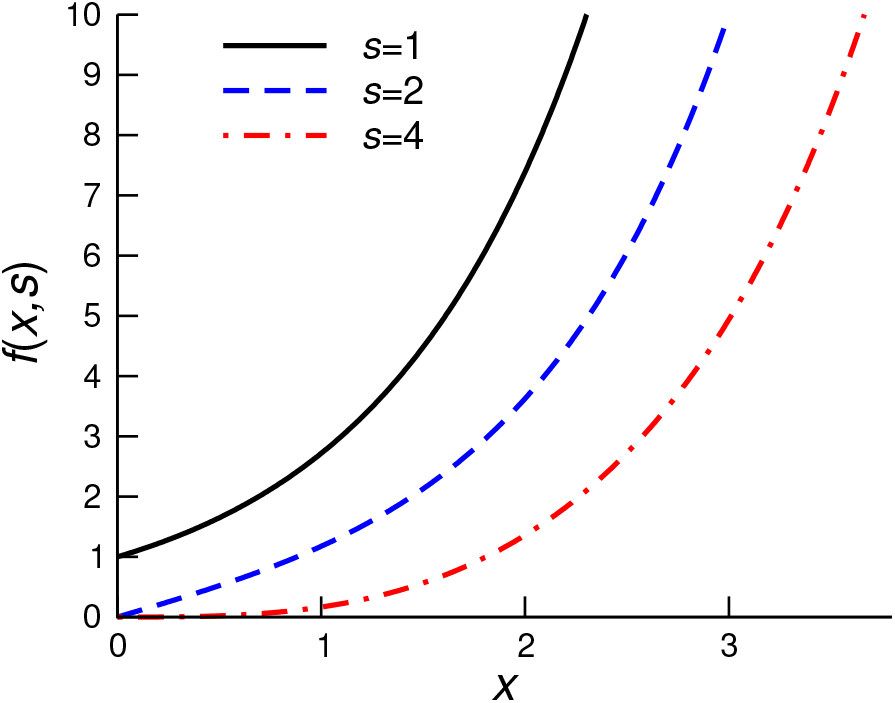
Shape of *f* (*x, s*) for different values of *s*.

When *s* is a natural number

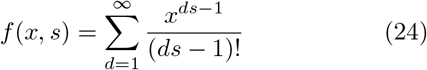

is a subseries of the exponential function. This series has been calculated [13, 14], resulting in

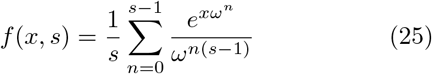

where *ω* = *e*^2*πi/s*^. For *x ≫* 1, since *ℜ*(*ω*^*n*^) *<* 1 for all *n* = 1, …, *s −* 1, we obtain

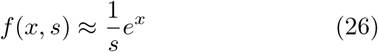

Based on this asymptotic behaviour, for *R*^1*/s*^*γt ≫* 1 equation (18) is approximated by

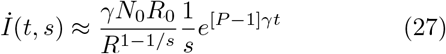

Where

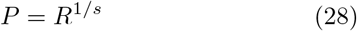

is the population reproductive number. Equation (28) coincides with the result from Wallinga and Lipsitch based on the Lotka-Euler equation [15].

The equation for the population reproductive number (28) is a tool to estimate the basic reproductive number from empirical data for the generation interval distribution and the doubling time. According to equation (27), the disease doubling time is given by

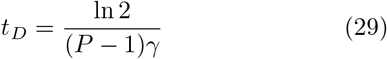

*t*_*D*_ is estimated from the plot of the number of new cases as a function of time. *s* and *γ* are estimated from a fit to the generation interval data. Then, using equations (28) and (29) we can estimate *P* and *R*.

Equation (28) is definitive proof that the shape of the generation interval distribution determines the relationship between the basic (*R*) and population (*P*) reproductive numbers. For *s* = 1 we recover the SIR model equation (4), when the individual and population reproductive numbers coincide. For *s* = 2 we recover the case *α* = *β* of the SEIR model in equation (9). Figure 3 shows the population reproductive number as a function of *s* for two different local reproductive numbers. When *R >* 1, *P* decreases monotonically with increasing *s*. When *R <* 1, *P* increases monotonically with increasing *s*. In either case, *P* approaches 1 for large *s*. Note, however, that the shape parameter *s* does not change the fact that if *R >* 1 then *İ*(*t*) grows exponentially, while *I*(*t*) decays exponentially when *R <* 1.

**FIG. 3.**
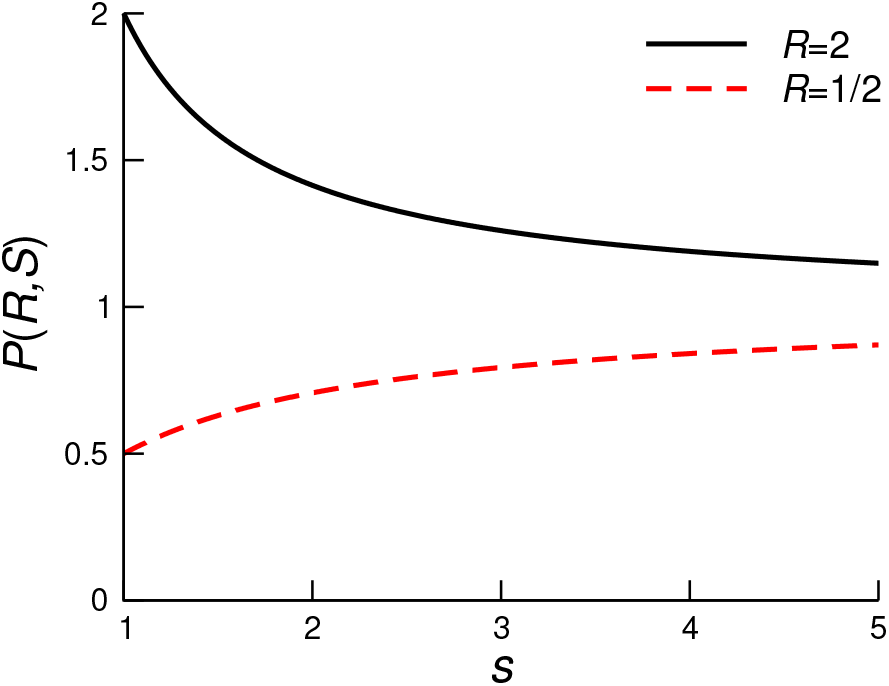
Population reproductive number *P* as a function of *s* for *R >* 1 (solid line) and *R <* 1 (dashed line).

The gamma distribution of generation intervals has a similar effect in the context of heterogenous mixing patterns between individuals according to types [16, 17]. The outcome is similar to equation (28), replacing *R* by the largest eigenvalue *ρ* of the mixing matrix of reproductive numbers (equation 12 in Ref [17]). After making this substitution, we obtain the population reproductive number for the multi-type generalization

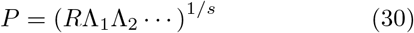

where Λ_*i*_ is the largest eigenvalue of the *i*th mixing matrix (*e*.*g*., age, mask use, etc) [17].

## VII. NUMERICAL SIMULATIONS

I have performed agent based simulations to test the relationship between the population and agent reproductive numbers, Eq. 28. The simulation steps were reported in Ref. [17]. Here, I present a concise description. The simulations take place in a virtual city. The virtual city is composed of a place-to-place network and the mobility of individuals (agents) through the network. Most of the city parameter are inspired on numerical simulation for the city of Portland [2], containing of the order of *n*_*a*_ = 1000000 individuals (agents) and *n*_*p*_ = 100000 places.

### Place-to-place network

The place-to-place network is modelled by a Barabási-Albert network [18]. Specifically, starting from a complete graph of *m*+1 nodes, new nodes are added one at the time up to *n*_*p*_ nodes. Each time a node is added, it is connected to *m* nodes in the pre-existing graph. The node to which each of the *m* edges is attached to is selected with a probability proportional to the node degree. The degree distribution of the Barabási-Albert network has a power-law tail with power-law exponent -3 [18].

### Agents mobility

Agents switch location at a rate *ω*_*i*_, *i* = 1, …, *n*_*a*_, where the *ω*_*i*_ are random variables with a gamma distribution Prob 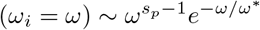, where *ω*^*∗*^ and *s*_*p*_ are the location and shape parameters, respectively. I will set *ω*^*∗*^ = 1 switch per day and *s*_*p*_ = 2, which gives a mode at 1 switch per day. This means that individuals will be in about two places per day, one where they started the day and the other where they switch to, as observed for the Portland simulation. Furthermore, the number of visitors at a given place is proportional to the number of neighbours, i.e. the degree in the place-to-place network. Since, the degree distribution of the Barabási-Albert network has a power law tail with exponent -3, the number of visitors to a place also has a power law tail with an exponent -3 [17], which is roughly what observed for Portland [2].

### Disease transmission

An infectious disease model is simulated in the virtual city introducing a patient zero and constraining the disease transmission to individuals at the same place. To model the disease dynamics within an individual, I introduce *s−*1 intermediate states and an infectious state. At a given place, I assume homogenous mixing and infected agents transmit the disease to susceptible agents at rate *ξ*. When a susceptible agent becomes infected he/she transits over *s −*1 exposed states, the infectious state and the removed state. The transition between these states is assumed state independent and at rate *γ. s* = 1 corresponds with the SIR model. For *s* = 2 the intermediate state is the exposed state and we recover the SEIR model with *α* = *γ*. For all *s* we obtain the gamma distribution of generation intervals (16). Here I will use *ξ* = 4 transmission attempts per day and *γ* = 1*/*3 per day. By transmission attempt I mean: there is an attempt of transmission from an infectious primary case to a potential secondary case at the location of the primary case, the potential secondary case is selected with uniform probability among all other agents at that location, but the transmission will happen if and only if the potential secondary case is in the susceptible state.

### Realizations

At each realization I generate a new place-to-place network, assign new switching rates to agents and run the disease transmission model. Averages are calculated over time intervals of 1 day and 100 realizations.

Figure 4A reports the average number of new infections as a function of time. The lines are fits to the exponential function in Eq. (27). The fit is restricted to the initial growth phase, selecting data points in the time interval 0 *< t <* 0.8*∗ t*_0_, where *t*_0_ is the time when *İ*(*t*) is maximum. Given that *γ* is known, from the fit to Eq. (28) we obtain an estimate of *P*, reported in Fig. 4B. The case *s* = 1 corresponds with the SIR model and *P* = *R*. In the fully mixed scenario, i.e. only one place, we would expect a reproductive number *R* = *ξ/γ* = 12. However, the network structure of the virtual city limits the homogeneous mixing to individuals within the same location. We obtain the smaller value *R≈* 7 *<* 12 (Fig. 4B, *s* = 1). Once *R* is known, we can use (28) to obtain a theoretical estimate of *P* for *s >* 1, which corresponds with the theoretical line in Fig. 4B. The agreement between the simulation symbols and the theoretical line is very good, validating the analytical calculations.

**FIG. 4.**
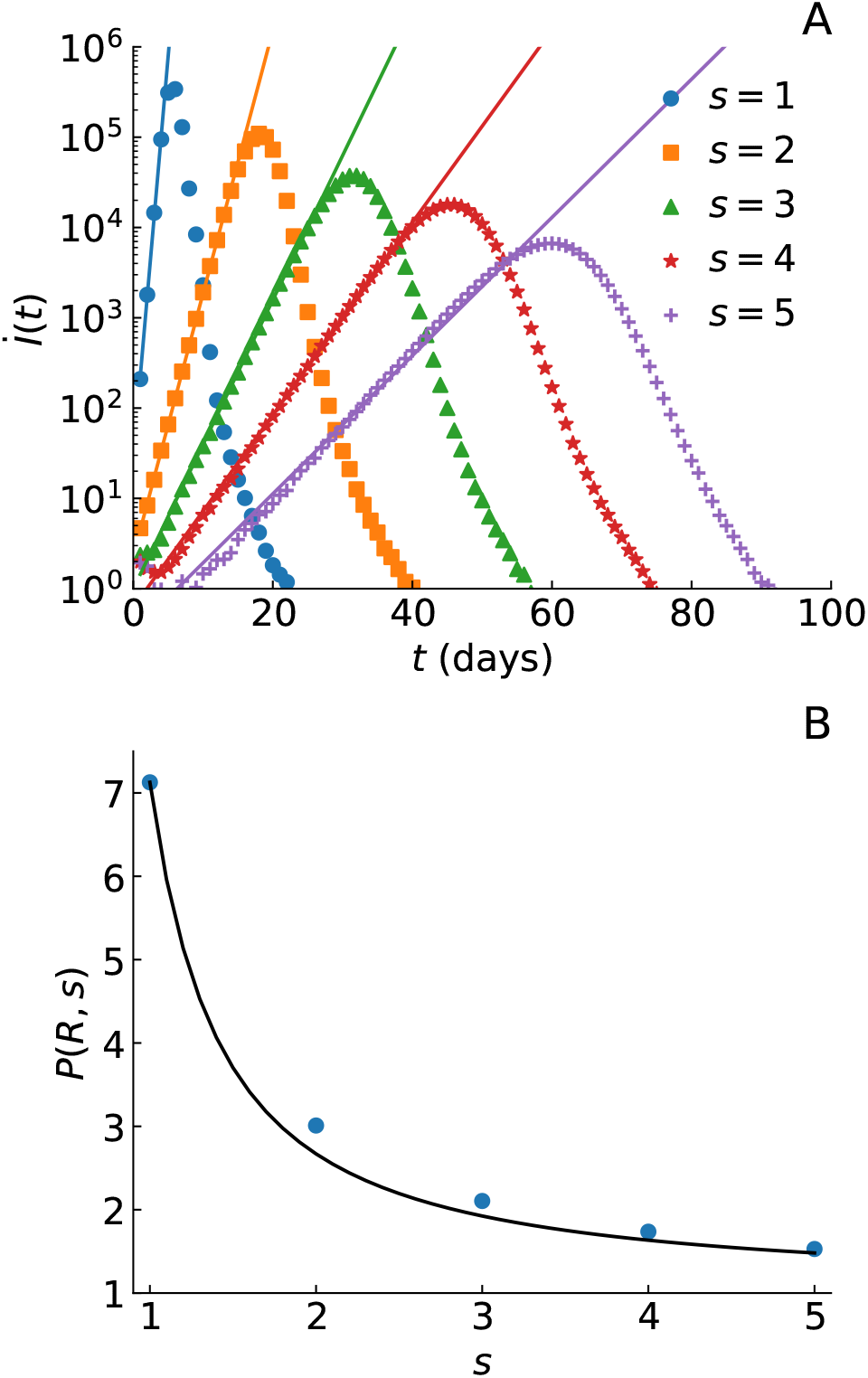
Numerical simulations of disease transmission in a virtual city. A) Average number of new infectious as a function of time for different number of intermediate states (symbols). The lines are fits to the exponential function in Eq. (27), providing an estimate of *P*. B) Population reproductive number *P* as a function of *s*, as obtained from the fitting to the numerical data (symbols) and from the expected theoretical line (28) (line).

## VIII. CONCLUSIONS

In conclusion, the branching process formalism allows for a flexible description of infectious disease outbreaks that can be fully based on empirical distributions. At short times the outbreak grows as a power law, *t*^*s−*1^,where the exponent is determined by the shape parameter of the gamma distribution. Therefore a power law growth is not incompatible with the homogeneous mixing approximation as previously claimed [19]. Furthermore, this power law should not be confused with the long-time power law induced by the truncation of the disease transmission at a maximum generation, as expected from an imposed lockdown for example [8, 10].

## Data Availability

All data are contained within the submission.

## References

[1] F. Liljeros, C. R. Edling, L. A. Amaral, H. E. Stanley, and Y. Aberg, Nature 411, 907 (2001).

[2] S. Eubank, H. Guclu, V. S. Kumar, M. V. Marathe, A. Srinivasan, Z. Toroczkai, and N. Wang, Nature 429, 180 (2004).

[3] R. Pastor-Satorras and A. Vespignani, Phys. Rev. Lett. 86, 3200 (2001).

[4] M. E. J. Newman, Phys. Rev. E 66, 016128 (2002).

[5] M. Boguñá, R. Pastor-Satorras, and A. Vespignani, Phys. Rev. Lett. 90, 028701 (2003).

[6] A. Vazquez and Y. Moreno, Phys. Rev. E 67, 015101 (2003).

[7] M. Keeling, P. Rohani, and P. U. Press, Modeling Infectious Diseases in Humans and Animals (Princeton University Press, 2008).

[8] A. Vazquez, Phys. Rev. Lett. 96, 038702 (2006).

[9] A. Vazquez, “Causal tree of disease transmission and the spreading of infectious diseases,” in Discrete Methods in Epidemiology, DIMACS Series in Discrete Mathematics and Theoretical Computer Science, edited by J. Albello and G. Cormode (2006) pp. 161–176.

[10] A. Vazquez, Phys. Rev. E 102, 040302 (2020).

[11] K. Sun and et al, medRxiv 2020.08.09.20171132 (2020).

[12] Z. Du, X. Xu, Y. Wu, L. Wang, B. Cowling, and L. A. Meyers, Emerging Infectious Disease journal 26, 1341 (2020).

[13] L. Rubel and K. Stolarsky, The American Mathematical Monthly 87, 371 (1980).

[14] H. Chen, International Journal of Mathematical Education in Science and Technology 41, 538 (2010).

[15] J. Wallinga and M. Lipsitch, Proceedings of the Royal Society B: Biological Sciences 274, 599 (2007).

[16] A. Vazquez, Phys. Rev. E 74, 066114 (2006).

[17] A. Vazquez, medRxiv (2020), 10.1101/2020.10.09.20210252.

[18] A.-L. Barabási and R. Albert, Science 286, 509 (1999).

[19] S. Thurner, P. Klimek, and R. Hanel, Proceedings of the National Academy of Sciences 117, 22684 (2020).

